# Using a transection paradigm to enhance the repair mechanisms of an investigational human cell therapy

**DOI:** 10.1101/2022.02.22.22271022

**Authors:** Monica J. Chau, Jorge E. Quintero, Paula V. Monje, S. Randal Voss, Andrew S. Welleford, Greg A. Gerhardt, Craig van Horne

## Abstract

One promising strategy in cell therapies for Parkinson’s Disease (PD) is to harness a patient’s own cells to provide neuroprotection in areas of the brain affected by neurodegeneration. No treatment exists to replace cells in the brain. Thus, our goal has been to support sick neurons and slow neurodegeneration by transplanting living repair tissue from the peripheral nervous system into the substantia nigra of those with PD. Our group has pioneered the transplantation of transection-activated sural nerve fascicles into the brain of human subjects with PD. Our experience in sural nerve transplantation through FDA-regulated clinical trials has supported the safety and feasibility of this approach. We are among the first to collect human sural nerve both before and after transection and to perform single nuclei RNA sequencing to determine the cell types present. We collected nerve tissue before and approximately 2 weeks after sural nerve transection for immunoassays from 15 participants, and collected from two additional participants for single nuclei RNA sequencing. We quantified the expression of key neuroprotective and anti-apoptotic genes along with their corresponding protein levels using immunoassays. The single nuclei data clustered into 10 distinctive groups defined on the basis of previously published cell type-specific genes. Transection-induced reparative peripheral nerve tissue showed RNA expression of neuroprotective factors and anti-apoptotic factors across multiple cell types after nerve injury induction. Key proteins of interest (*BDNF, GDNF, beta-NGF, PDGFB, and VEGF*) were significantly upregulated in reparative tissue compared to naïve. These results provide insight on this repair tissue’s utility as a neuroprotective cell therapy.

**Clinical Trial:** Clinicaltrials.gov (Trial registration number NCT02369003)

https://clinicaltrials.gov/ct2/show/NCT02369003).

## Introduction

The use of human embryonic and neural stem cells have limitations as cell therapies for Parkinson’s Disease (PD)^1-3^. Obtaining embryonic or fetal cells can be ethically challenging, and embryos for transplantation are not always readily available^1,2,4^. Furthermore, they are not autologous tissues and require the patient to use immunosuppressing drugs. Other stem cell sources include autologous induced pluripotent stem (iPS) cells differentiated into dopaminergic progenitors; however their use in clinical testing in PD is still in its infancy^5^. Further, incompletely reprogrammed cells can elicit harmful immune responses^6,7^. A more feasible approach could be to use the body’s own repair mechanisms. Autologous tissue, like peripheral nerve has robust repair capabilities, is readily available, and can be efficiently procured^8,9^. Our strategy is to harness the patient’s own reparative peripheral nerve tissue and implant it to provide neuroprotection to areas of the brain affected by neurodegenerative disease.

After injury, cells in the peripheral nervous system (PNS) undergo a highly orchestrated transformation to regenerate and re-establish function to the extremities^10-12^. As part of an investigational cell therapy that we are currently trialing, we implanted active reparative peripheral nerve tissue into the substantia nigra of participants with PD. No treatment exists to repair damaged brain cells^8^, thus our goal was to slow neurodegeneration by implanting reparative living cells into the substantia nigra^5,9,13^.

Cell therapy strategies can affect disease progression either by replacing dead or dying cells, or by promoting cell-survival and neuroprotection via secretion of paracrine and neurotrophic factors. Implanting peripheral nerve tissue would not replace sick neurons of the central nervous system, but could provide neuroprotective, anti-inflammatory, anti-apoptotic, and pro-regenerative factors to support dying cells^14-16^. Furthermore, implanting autologous peripheral nerve tissue has the major advantages of being readily obtainable from patients and circumventing host immune rejection. If obtained from a sensory nerve like the sural nerve, the side effects, if any are mostly paresthesias and hypoesthesia in cutaneous distribution of the nerve^17,18^. In our previous studies, participants have reported that these incidents were not bothersome in the long-term^9,13,15^.

An ideal cell therapy against neurodegeneration would be robust enough to slow disease progression by providing neuroprotective, anti-apoptotic, and anti-inflammatory support to unhealthy cells while maintaining its potency from harvesting through deployment. Our approach has been to implant a supportive milieu of neuroprotective factors with the expectation that a combination of factors is more durable and effective than using a single neuroprotective factor therapy^19,20^. To date, definitive trials of single-neuroprotective factor therapy, *e*.*g*. glial-cell derived neurotrophic factor (GDNF), have shown mixed results in slowing PD progression^21,22,19,23^. The survival of neural stem cells after transplantation is limited since a proportion of cells die within days after transplantation into the brain^24-26^. Anti-apoptotic factors in the implanted cells may bolster cell survival after implantation^27^.

Even though there is a great deal known about the reparative microenvironment of injured peripheral nerves in animal models, data on human peripheral nerve repair and its neuroprotective properties is sparse. To fill this gap in knowledge, we collected human peripheral (sural) nerve tissue before and after transection injury (herein referred to as naïve and reparative for consistency). We used single nuclei RNA sequencing, and immunoassays to generate a profile of the cell types, the RNA expression, and protein content of key neuroprotective factors present in reparative peripheral nerve tissue. The objective of this study is to report our findings of the specific cell types and contents of the reparative human nerve. Our results focus primarily on the properties and contents of the reparative tissue as this is the final product transplanted into the brain. This information will provide insight on this tissue’s utility as a neuroprotective cell therapy.

## Methods

### Research participants

The collection of peripheral nerve tissue was approved as part of a more expansive clinical trial that received approval from the University of Kentucky’s Institutional Review Board and was registered at clinicaltrials.gov (NCT02369003). The participants provided written informed consent. Peripheral nerve tissue of the sural nerve was collected from 15 participants before and after sural nerve transection *in situ* for immunoassay studies – the range of differences between naïve and reparative tissue has been previously published by Chau et al^15^. Sural nerve tissue was collected from two additional participants for single nuclei RNA sequencing studies.

Peripheral nerve tissue samples were collected from 15 participants for immunoassays (Mean 60 years old, range 51 - 69 years, assigned birth sex: 9 male/6 female, years since PD diagnosis mean 10 years, range 4-17 years). Samples were collected from two additional participants for single nuclei RNA seq (ages were in the range of 46-50 and 56-60 years old, assigned birth sex: both male, years diagnosed with PD: range 5-10 years). The time between transection of the nerve (naïve) and collection of regenerating peripheral nerve tissue was 12 and 17 days for participants 1 and 2.

### Peripheral nerve transection and tissue collection

Transection of the peripheral nerve and the tissue collection have been previously described^8,9,13^. Figure 1 illustrates the naïve and transection injury-induced sural nerve collection approximately 2 weeks after transection. Conventionally, naïve nerve tissue is defined as tissue that had not degenerated. Our naïve tissue had been transected (and flash-frozen after) in order to remove it from the body, so it is not completely uninjured. However, for the purposes of this study and ease of understanding, we will call this “naïve” tissue at 0 days relative to the “reparative” tissue at approximately 2 weeks.

**Figure 1.**
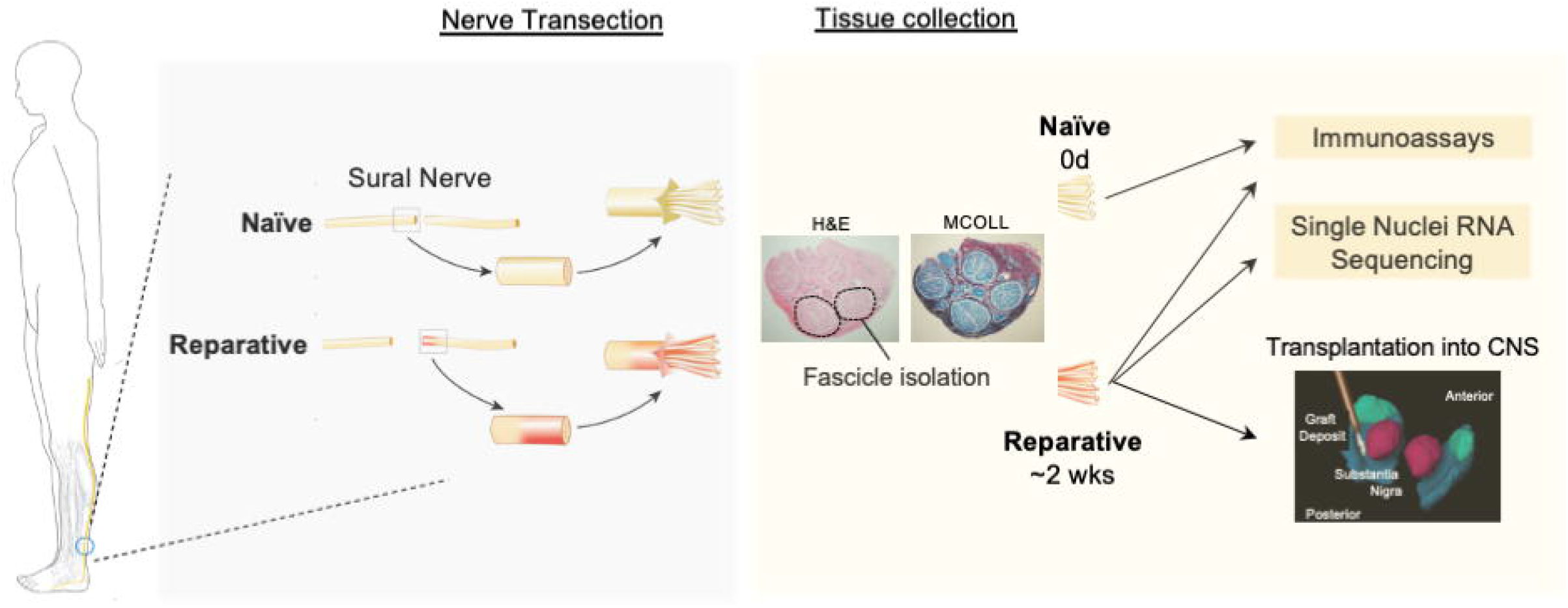
Study Overview. This overview illustrates our sural nerve transection approach and subsequent tissue collection of the naïve and reparative nerve tissues. One to two centimeters of nerve was excised, which we called “naïve” nerve tissue. Approximately 14 days after, one to two centimeters from the distal nerve stump of the same nerve was excised, which we called “reparative” nerve tissue. Cross-sections of reparative sural nerve was stained with H&E (left) and Luxol fast blue (LBF)/MCOLL staining to show myelin and collagen. Individual nerve fascicles were separated, snap-frozen, and used for single nuclei RNA seq and immunoassays or implanted directly into the brain as part of our clinical trial. Shown here is a 3D view of reparative nerve fascicles implanted into the substantia nigra in our cell therapy trial.

Briefly, the neurosurgeon identified the neurovascular bundle containing the sural nerve in the ankle and transected it and removed 1-2 cm of nerve of naïve tissue (Fig. 1). The nerve tissue was cleaned of loosely external tissues (fat tissue, blood vessels), and individual nerve fascicles (usually 6-10 per patient) were separated manually. The fascicles were cleaned of adherent connective tissue and snap-frozen in centrifuge tubes on crushed dry ice. The fascicles were stored in a -80 Freezer for analysis by immunoassays (as in Welleford et al.^13^). Approximately 2 weeks later, for both analysis and implantation into the substantia nigra for the clinical trial (Fig 1), the ankle incision was reopened and 1-2 cm of the injured peripheral nerve tissue was excised from the distal nerve stump. The individual nerve fascicles were separated, snap-frozen, and stored for assays as described above for the naive tissue^13^. Transection-injured tissue collection time from dissection to snap-freezing was 32 minutes for one participant, and 67 minutes for the other participant. These frozen samples of nerve fascicles were used in single nuclei RNA seq and immunoassays (weight per sample ranged from 40 to 170 mg).

### Histology

Reparative peripheral nerve was stained with H&E and MCOLL staining. MCOLL staining distinguishes myelin, collagen fibers, and cells in the peripheral nerve^28^. The nerve was placed in 4% paraformaldehyde solution, then embedded in paraffin blocks for histology^29^. Sections were all taken from within approximately 1 mm from the end of each nerve (since the sections were cut from the terminal end).

### Single nuclei RNA-Seq

Single nuclei RNA sequencing and analysis were conducted by Singulomics Corporation (https://singulomics.com, Bronx NY). In summary, frozen sural nerve fascicles (1-2 cm each) were homogenized and lysed with Triton X-100 in RNase-free water for nuclei isolation. The isolated nuclei were purified, centrifuged, and resuspended in PBS with BSA and RNAse Inhibitor. The nuclei were diluted to 700 nuclei/ul and loaded to 10x Genomics Chromium Controller to encapsulate single nuclei into droplet emulsions following the manufacturer’s recommendations (Pleasanton, CA, United States). Library preparation was performed according to the instructions in the Chromium Next GEM 3’ Single Cell Reagent kits v3.1. Amplified cDNAs and the libraries were measured by Qubit dsDNA HS assay (Thermo Fisher Scientific, Wilmington, DE) and quality assessed by BioAnalyzer (Agilent Technologies, Santa Clara, CA). Libraries were sequenced on a NovaSeq 6000 instrument (Illumina, San Diego, CA, United States), and reads were subsequently processed using 10x Genomics Cell Ranger analytical pipeline (v5.0) and human GRCh38 reference genome with introns included in the analysis. Dataset aggregation was performed using the cellranger aggr function normalizing for the total number of confidently mapped reads across libraries.

Seurat 4.0.1 was used to further clean and normalize the data. The data from barcodes with mitochondrial genes at a level of <5% of total gene counts and with a minimum of 1400 UMI counts were retained. Gene read counts were normalized with the Seurat ‘NormalizeData’ function. The top 3000 highly variable genes were identified using Seurat ‘FindVariableFeatures’ function, which were used for principal component analysis using Seurat ‘RunPCA’ function. Clustering was done using Seurat ‘FindClusters’ function based on 11 PCAs. The ElbowPlot test was done to determine the number of PCAs used. Visualization of the cells was performed using the Uniform Manifold Approximation and Projection for Dimension Reduction (UMAP) algorithm as implemented by the Seurat ‘runUMAP’ function. Violin plots were graphed as a log2 fold average count for RNA expression in each defined cell cluster through C Loupe 5.0 software (10x Genomics, Pleasanton, CA).

### Immunoassays for neuroprotective and anti-apoptotic factors

To quantify the neuroprotective and anti-apoptotic factors of interest present in reparative nerve, tissues were analyzed using enzyme-linked immunosorbent assay (ELISA and multiplex Luminex® immunoassays (Cincinnati Children’s hospital flow cytometry core). We analyzed reparative tissues from 15 participants.

Analyte concentrations in the regenerating nerve tissue sample supernatants were determined by ELISA according manufacturer’s protocol. The sources and dilutions used were: Neuroprotective factors: cerebral dopamine neurotrophic factor (CDNF) (Abcam, Waltham, MA), tissue samples were diluted 1:10. Nerve Growth Factor Receptor (NGFR) (ThermoFisher Scientific, Carlsbad, CA), tissue samples were diluted 1:2. EPO concentrations in the sample supernatants were determined by using MilliplexTM Multiplex kits (MilliporeSigma, Darmstadt, Germany) and brain derived neurotrophic factor (BDNF), nerve growth factor (NGF, beta-NGF), Platelet-Derived Growth Factor-AA (PDGF-AA) -BB (PDGF-BB) -AB (PDGF-AB), vascular endothelial growth factor (VEGF), glial cell-derived neurotrophic factor (GFAP, and neurotrophin-3 (NT-3) were determined by Human Magnetic Luminex Assays (R&D Systems, Minneapolis, MN) according to the manufacturer’s protocol. Anti-apoptosis factors: Nuclear factor erythroid 2-related factor 2 (NRF2) (ThermoFisher Scientific, Carlsbad, CA), tissue samples diluted to 1:2. B cell lymphoma 6 (BCL-6) (MyBiosource, San Diego, CA), tissue samples were neat.

### Statistical analysis

For each protein analyzed with immunoassay, a paired t-test (naïve paired with reparative matched control from the same participant) was used to determine statistical significance (p<0.05).

### Availability of Data and Materials

Data files (.cloupe) can be obtained from the UKnowledge database. (https://uknowledge.uky.edu/).

## Results

### Reparative peripheral nerve tissue contains transcriptionally distinctive cell types

We used single nuclei RNA seq to identify the cell types present in reparative peripheral nerve tissue based on transcriptional profiling (Fig. 2A). Single nuclei RNA Seq data from two participants were aggregated (Fig. 2A). Cell clustering by cell type was reproducible across both participants (Fig. 2B). The data clustered into 10 cell groups and were defined based on characteristic genes in previously published single cell data for each particular cell type (Fig. 2C)^30-39^. Cell counts were obtained from the single nuclei RNA Seq analysis and each cluster’s percentage was reported (Fig. 2D). An output of 2425 cells and 2643 cells were used from both participants.

**Figure 2.**
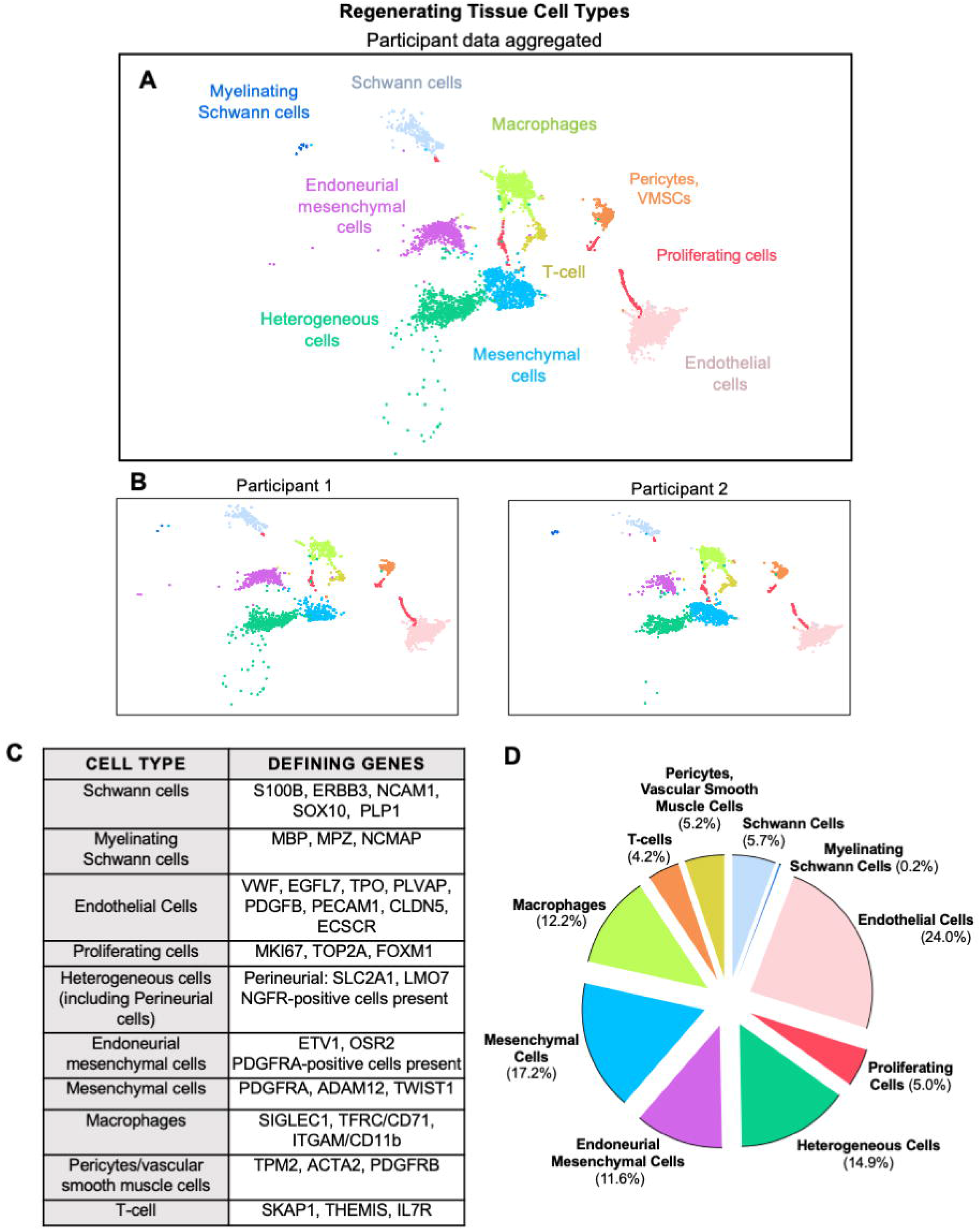
Regenerating peripheral nerve tissue contains several cell types including regenerating cells. **A**. Aggregate of single nuclei RNA seq from two participants. Ten unique cell type clusters were present in the reparative tissue. **B**. Data from two participants show similarity in their cellular profiles. **C**. The data clustered into 10 cell groups and were defined based on characteristic genes previously published for each particular cell type. **D**. Each cell cluster’s percentage of total number of cells is reported.

Cell clusters observed included typical Schwann cells (defined by their expression of *S100B, ERBB3* and other lineage-specific genes), macrophages (*ITGAM*), T-cells (*SKAP1*) endothelial cells (*PECAM1*), and pericytes (*ACTA2*) (Fig. 2C). The content of mature (myelinating) Schwann cells was negligible (0.2%). The small number of (myelinating) Schwann cells was an expected finding considering that the nerve transection had a removal of the intermediate segment. The proximal and distal stumps could not physically reconnect, thus could not form axonal regeneration. A proportion of Schwann cells, macrophages, endothelial cells, and T-cells exhibited expression of genes associated with active cell division (Fig. 2A, B, red clusters). The distribution of proliferative cells was coincidentally conserved in the two donors (Fig. 2B).

Different than what we had expected, a large proportion of the cells were mesenchymal cells including a cluster of mesenchymal cells expressing stem cell-associated genes (*TWIST1*^*35*^, *PDGFRA*^*35*^, and *ADAM12)* and endoneurial mesenchymal cells (Fig. 2C, D). These endoneurial mesenchymal cells were defined by *ETV1*^30^,^38^, and *OSR2*^31^,^38^,^39^, and were positive for the mesenchymal marker, *PDGFRA* (Fig. 2C). Another unexpected result was that one cell cluster contained a heterogenous mix of cells including perineurial cells with the expression of markers *SLC2A1*^30^ and *LMO7*^40^, and possibly repair Schwann cells (*NGFR*)^35^.

### Reparative peripheral nerve tissue shows RNA expression of neuroprotective factors 2 weeks after nerve transection

In this study, we aimed to identify a select group of neuroprotective factors in the regenerating tissue and localize the RNA expression to specific cell types (Fig. 3). This differs from previous our work in the whole nerve tissue in which Welleford et al. had identified gene expression of neuroprotective and anti-apoptotic factor pathways^13^. UMAP plots illustrate the cell-type expression of the neuroprotective factors of interest: *BDNF, EPO, CDNF, GDNF, NGF, PDGFA, PDGFB, EPO*, and *VEGFA* (Fig. 3A). The accompanying violin plots show the relative RNA expression level (log2 average) and frequencies for each cell type in the regenerating peripheral nerve tissue (Fig. 3A).

**Figure 3.**
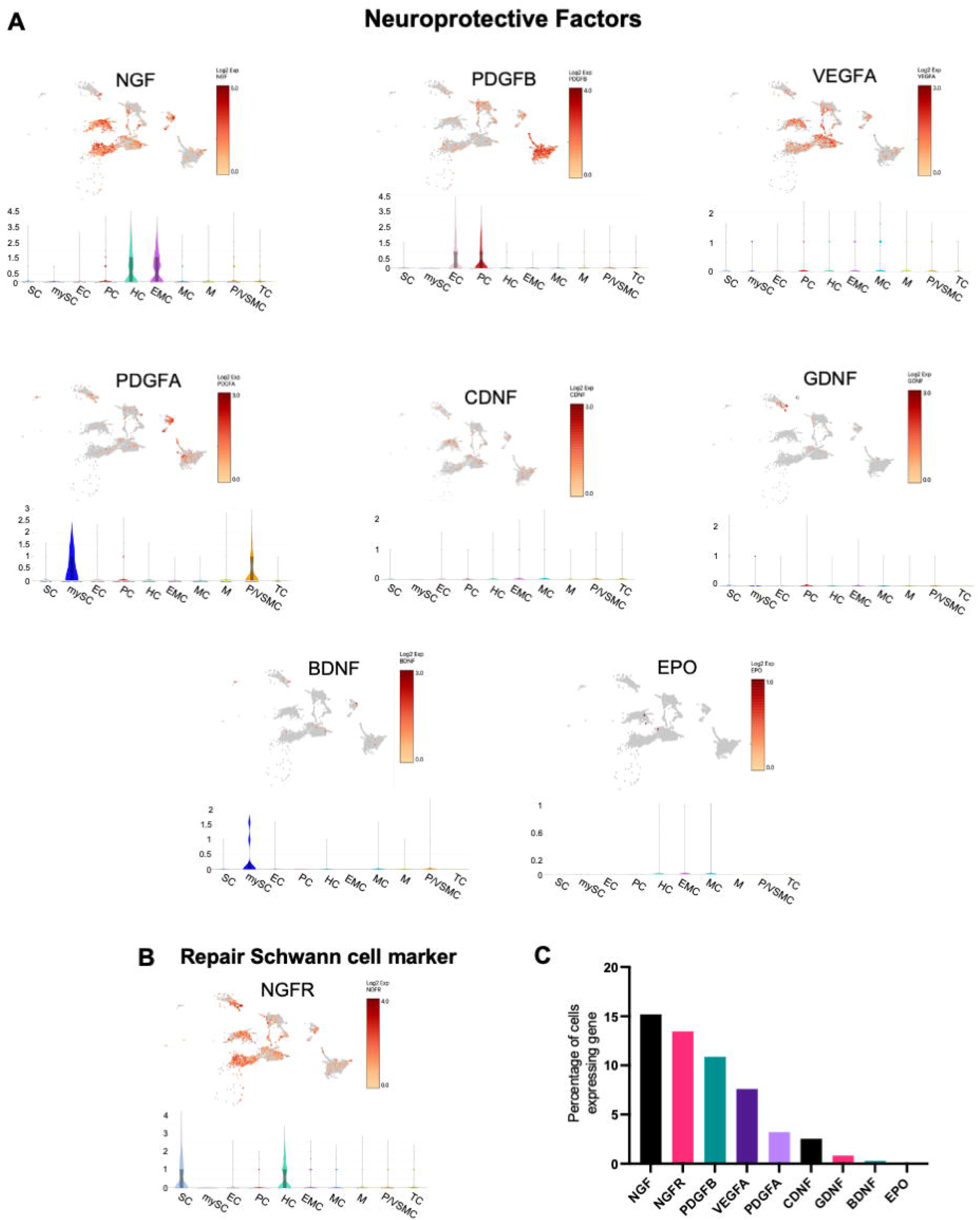
Reparative peripheral nerve tissue shows RNA expression of neuroprotective factors 2 weeks after nerve transection. **A**. Single nuclei RNA seq UMAP plots show cell-type expression of the neuroprotective factors of interest: *NGF, PDGFB, VEGFA, PDGFA, CDNF, GDNF, BDNF*, and *EPO*. The accompanying violin plots show the relative RNA expression level (log2 average) and frequencies for each cell type in the regenerating peripheral nerve tissue. *NGF, PDGFA, PDGFB*, and *VEGF* were localized to more than one cell type. *NGF* was localized to the heterogenous cell cluster and endoneurial mesenchymal cells, *NGFR* was localized to Schwann cells and the heterogenous cell cluster, *PDGFA* was localized to myelinating Schwann cells and pericytes/vascular smooth muscle cells, *PDGFB* was highly expressed in endothelial cells. **Cell type key:** SC-Schwann cell, mySC-Myelinating Schwann Cells, EC-Endothelial cells, PC-proliferating cells, HC-Heterogeneous cells, EMC-Endoneurial mesenchymal cells, MC-Mesenchymal cells, M-Macrophages, PC/VMSC-pericytes/vascular smooth muscle cells, TC-T-cells **B**. Repair Schwann cells are a source of neuroprotective factor release. *NGFR* is an abundantly expressed repair Schwann cell marker was found in the Schwann and heterogenous cell clusters **C**. Percentage of total cells expressing each key neuroprotective factors: *NGF* (15.2% of cells), *NGFR* (13.5% of cells), and *PDGFB* (10.9% of cells) were the most widely expressed factor across cells of the reparative peripheral nerve tissue while *GDNF* (0.8%), *BDNF* (0.3%), and *EPO* (0.04%) were the most limited in expression. Note that the x-axis maximum is 20%.

Even though certain factors (*GDNF, CDNF, EPO*) do not show very much RNA expression, we felt that it was important to include this data since we will compare them to their protein level expression.

Repair Schwann cells are a source of neuroprotective factor release in reparative nerve. We included the most abundant repair Schwann cell marker expressed in our tissue, *NGFR*^35^ to understand the relative location and presence of repair Schwann cells in our tissue (Fig. 3B).

Factors such as *NGF, PDGFA, PDGFB*, and *VEGF* were localized to more than one cell type (Fig. 3A, violin plots). *NGF* was localized to the heterogenous cells cluster and endoneurial mesenchymal cell cluster, *PDGFA* was localized to myelinating Schwann cells and pericytes/vascular smooth muscle cells, *PDGFB* was highly expressed in endothelial cells and proliferating cells. Among the key neuroprotective factors, *NGF* (15.2% of cells), and *PDGFB* (10.9% of cells) were the most widely expressed factor (RNA) across cells of the reparative nerve tissue while *GDNF* (0.8%), *BDNF* (0.3%), and *EPO* (0.04%) were the most limited in expression (Fig. 3B). NGFR was expressed in 13.5% of cells (Fig. 3B, note that the x-axis maximum is 20%).

### Reparative peripheral nerve tissue shows RNA expression of anti-apoptotic factors across multiple cell types

The anti-apoptotic factors, *NFE2L2 (NRF2), BCL2, BCL2L1 (Bcl-xl)*, and *MCL1* were expressed broadly and robustly in many of the cell types. Violin plots show the relative expression across cell types (log2 average) (Fig. 4A). Among the anti-apoptosis factors, *NFE2L2* (36.3% of cells), *BCL2* (28.6% of cells) were the most widely expressed factor across cells of the reparative peripheral nerve tissue while *BCL6* (16.9%) and *MCL1* (14.5%) were the most limited in expression. (Fig. 4B, note that the x-axis maximum is 40%).

**Figure 4.**
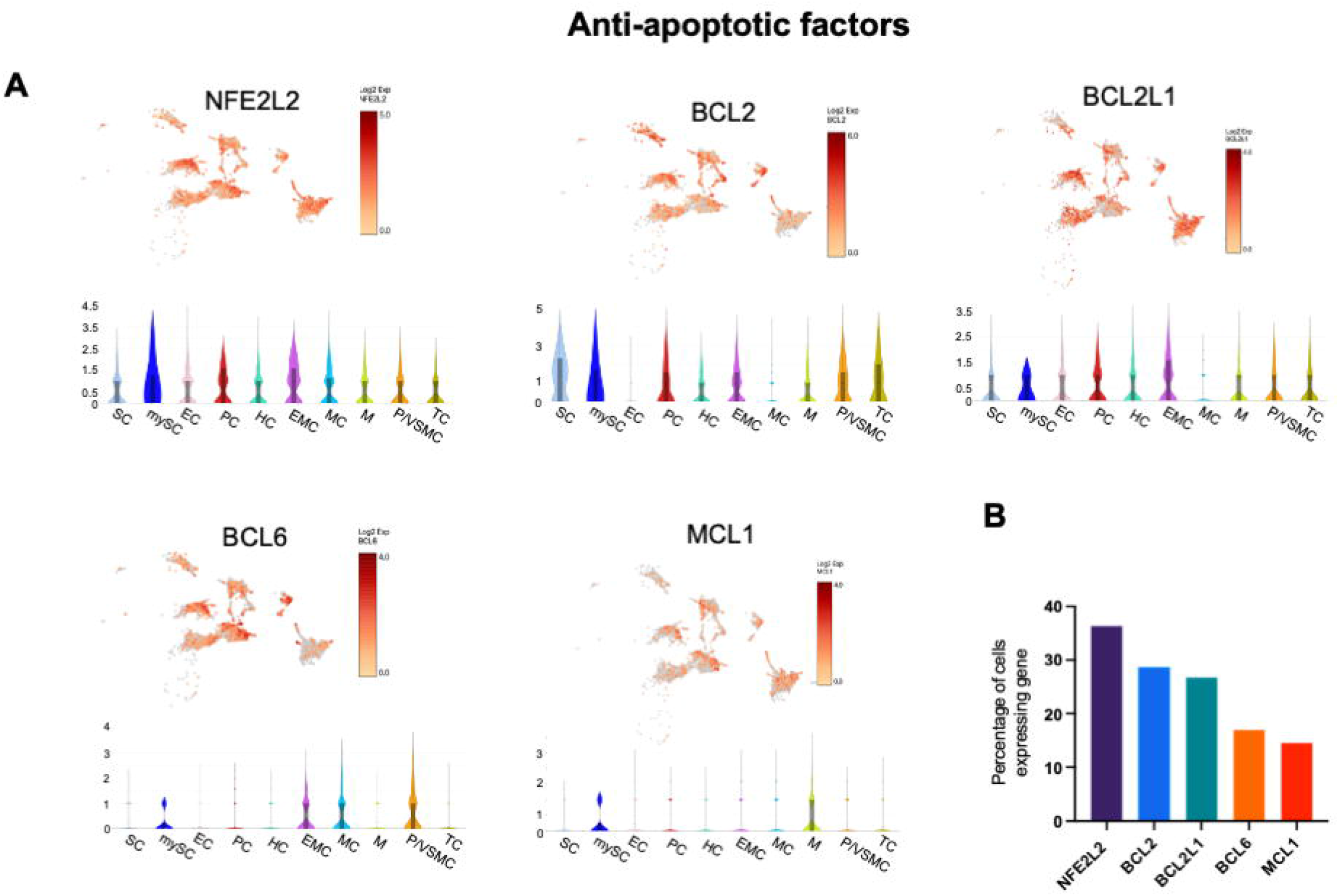
Reparative peripheral nerve tissue shows RNA expression of anti-apoptotic factors across multiple cell types. **A**. Anti-apoptosis factors were expressed broadly and robustly in many of the cell types of the reparative nerve for factors of interest: *NFE2L2 (NRF2), BCL2, BCL2L1 (Bcl-xl), BCL6*, and *MCL1* of reparative peripheral nerve tissue. Violin plots show the relative expression across cell types (log2 average). **Cell type key:** SC-Schwann cell, mySC-Myelinating Schwann Cells, EC-Endothelial cells, PC-Proliferating cells, HC-Heterogeneous cells, EMC-Endoneurial mesenchymal cells, MC-Mesenchymal cells, M-Macrophages, PC/VMSC-pericytes/vascular smooth muscle cells TC-T-cells **B**. Percentage of total cells expressing each anti-apoptotic factors: *NFE2L2* (36.3% of cells), *BCL2* (28.6% of cells) were the most widely expressed factor across cells of the reparative peripheral nerve tissue while *BCL6* (16.9%) and *MCL1* (14.5%) were the most limited in expression. Note that the x-axis maximum is 40%.

### Protein content of neuroprotective factors and anti-apoptotic factors

To measure the protein content of neuroprotective and anti-apoptotic factors in regenerating peripheral nerve tissue, we conducted immunoassays for several genes that we had characterized with single nuclei RNA Seq (Fig. 2-4). The mean protein concentration (and SD) in reparative peripheral nerve tissue samples is summarized in Fig. 5A. Following nerve transection, we found a significant upregulation in the proteins of several neuroprotective factors and the repair Schwann cell marker NGFR, in reparative vs. naïve samples, including BDNF (t(14) = 2.35, p = 0.034), GDNF (t(6) = 3.80, p = 0.009), beta-NGF (t(11) = 2.62, p = 0.024), NGFR (t(13) = 2.59, p = 0.022), PDGFB (t(14) = 2.12, p = 0.052), VEGF (t(14) = 2.36, p = 0.033) (Fig. 5B). The transection-injury also significantly raised the levels of BCL6 in the reparative peripheral nerve tissue (t(14) = 3.51, p = 0.004 Fig. 5C).

**Figure 5.**
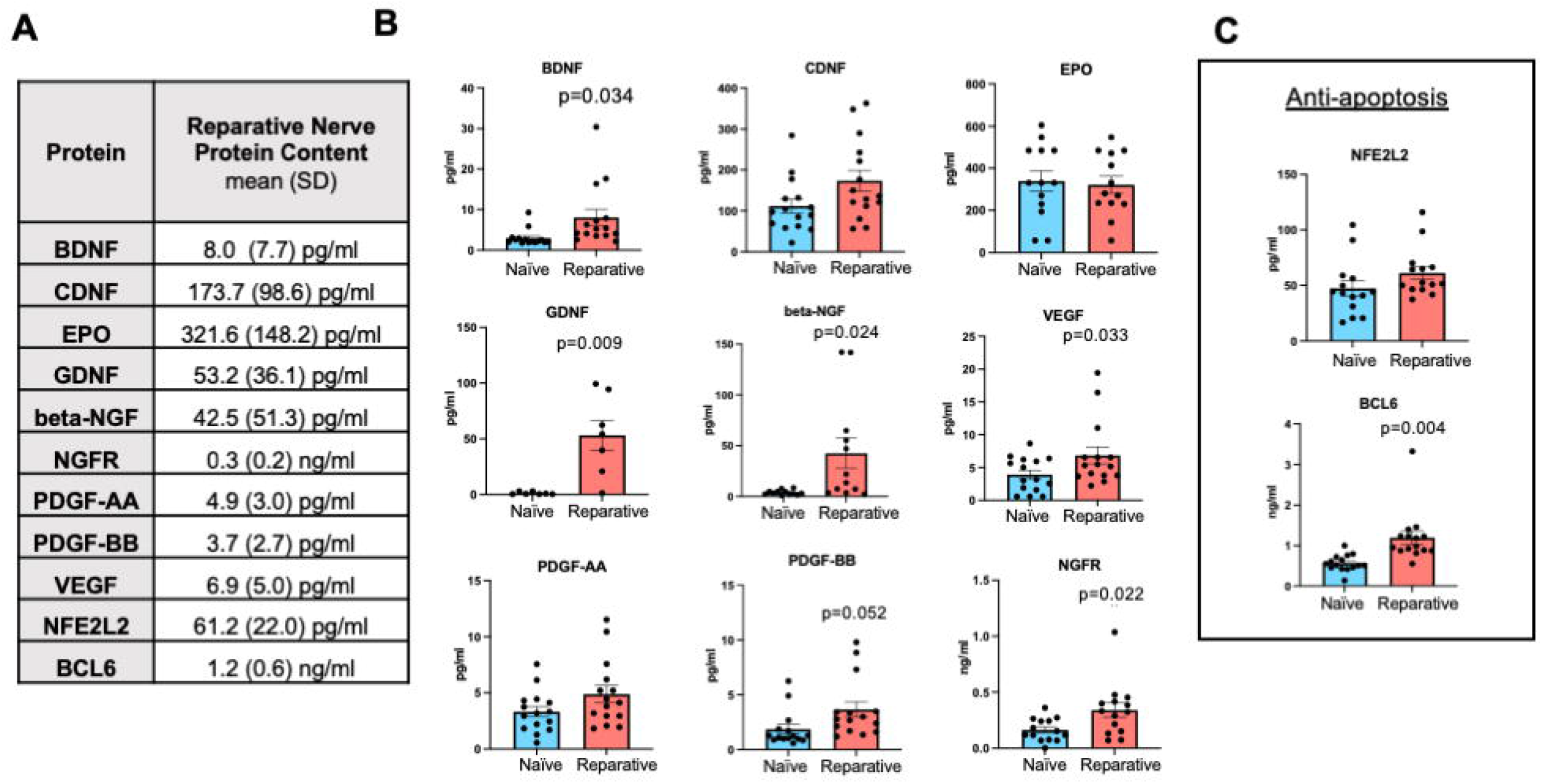
Protein content of neuroprotective factors and anti-apoptotic factors. **A**. The mean protein concentration (and SD) in regenerating peripheral nerve tissue samples is summarized. **B**. Following transection, there was significant upregulation in the proteins of several neuroprotective factors when comparing naïve with reparative tissue including BDNF (t(14) = 2.35, p = 0.034), GDNF (t(6) = 3.80, p = 0.009), beta-NGF (t(11) = 2.62, p = 0.024), PDGFB (t(14) = 2.12, p = 0.052), VEGF (t(14) = 2.36, p = 0.033). NGFR, a repair Schwann cell markers was also significantly upregulated (t(13) = 2.59, p = 0.022), **C**. After transection, reparative tissue also showed significantly increased levels of the anti-apoptotic factor, BCL6 in the regenerating peripheral nerve tissue (t(14) = 3.51, p = 0.004).

**Figure 6.**
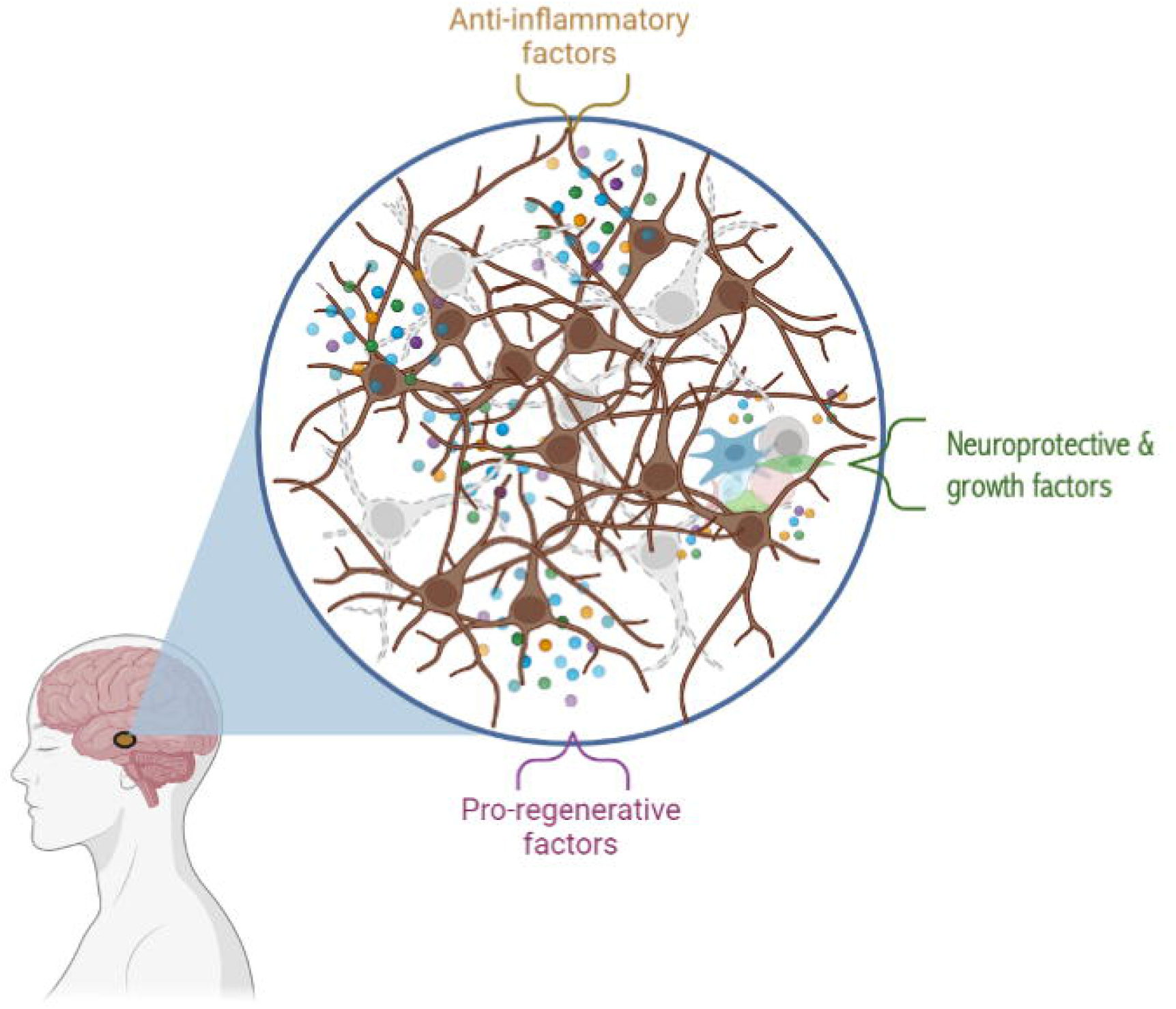
Proposed action of peripheral nerve transplant. Reparative peripheral nerve tissue transplanted into the substantia nigra of participants with PD may act in multi-factorial ways with paracrine effects on the surrounding tissue. Anti-apoptotic factors may contribute to cell transplant survival. Through this combination, a diversity of cell-types from regenerating peripheral nerve tissue could provide neuroprotective, pro-regenerative, and anti-inflammatory factors interacting with the degenerating cells in the CNS. Image created with BioRender.com

## Discussion

In this report, we focused on the final product that is implanted into the brain in our clinical trial, the reparative peripheral nerve. We identified the cell types found in the implanted peripheral nerve tissue. Further, this work details the distribution of neuroprotective and anti-apoptotic factors within these cell types, and their protein concentrations. In animal models, multiple cell types contribute to peripheral nerve repair notably repair Schwann cells^41-44^, endothelial cells, and immune cells such as macrophages^45-47^. In the two single nuclei RNA seq subjects with PD in this study, the major cell types in reparative human peripheral nerve tissue were consistent and reproducible (Fig. 2). Understanding the contents of this investigational cell therapy is a critical step in optimizing the product’s survival and its ability to neuroprotect vulnerable neurons. We collected same-subject matched naïve and reparative tissue to determine how transection injury affected the expression and cells. It is not typically feasible to collect tissue before an injury has occurred^48^, thus this type of information has not been available in humans before.

### Actively regenerating nerve tissue serves as a vehicle for neuroprotective factors

In this study, we show a clear upregulation of neuroprotective proteins. Interestingly, the results showed low mRNA expression for certain neuroprotective factors including *BDNF, GDNF, VEGF*, but the protein content was highly expressed. Low mRNA expression but high protein expression may mean that the mRNA was translated into protein at this time point.

To provide the putative beneficial factors found in peripheral nerve tissue requires the delivery of the collective content of peripheral nerve tissue and not a single cell type in isolation^49,50^. The combination of cell types has the benefit of including all of the neuroprotective, growth, pro-regenerative, cell survival factors, and anti-inflammatory factors, available from living reparative nerve tissue to support of degenerating cells in the CNS ^13,15^

The reparative human peripheral nerve is living tissue, as it is freshly dissected from the participant’s ankle and immediately implanted into the brain (Fig 1). For how long the neuroprotection continues after implantation remains unclear. We speculate that there is a persistent delivery of these factors up to a point to support degenerating cells based on the beneficial results that we have observed. In a previous report of participants that had received this implantation in a preliminary, open-label trial, we observed an improvement in Unified Parkinson′s Disease Rating Scale (UPDRS) part III motors scores at 12 months^9^. Further, post-mortem sections of a participant who had been engrafted with reparative peripheral nerve tissue to the midbrain 33 months earlier showed immunoreactivity to *NGFR* present in the area around the engraftment (unpublished). This suggests the presence of Schwann-like glia or repair Schwann cells^35^ which is a source of neuroprotective factor release and repair. Future studies should be designed to measure secretion levels from reparative peripheral nerve tissue to analyze this tissue beyond the implantation site.

Even though the anti-apoptotic factors that we highlight in this study are not secreted, their robust expression in the transplanted cells could bolster their own survival after implantation. It is typical to find high levels of anti-apoptotic genes in transected nerves in all cell types. These cells are primed for survival and reprogramming due to the transection^44,51,52^. We used this to our advantage to implant a robust product. A percentage of transplanted neural stem cells die due to the trauma and manipulation of cells days after transplantation into the brain^24-26^. This suggests that cell survival may be supported by a robust expression of anti-apoptotic factors in the implanted cells^27^. Additionally, neurotrophic factors are also inherently anti-apoptotic and pro-survival.

### Beneficial cell types in active reparative tissue

#### Repair Schwann cells

Much of the work surrounding the reparative cell types in peripheral nerve tissue focuses on repair Schwann cells^44,52,53^. After injury or transection, Schwann cells undergo an extensive reprogramming that transforms the mature myelinating and non-myelinating (Remak) cells into dedifferentiated, repair cells^44,52,53^. Repair Schwann cells release neuroprotective factors to facilitate axonal regeneration^44,53-55^. In the early response to injury, Schwann cells undergo an epithelial-mesenchymal transition (EMT)-like process in which cells become more similar to multipotent stem cells as evidenced by the upregulate the expression of stem cell-associated transcription factors such as Sox2, Notch1, and Oct6. This EMT-like process transforms cells to be similar to multipotent stem cells and release neurotrophic factors and support cell survival^34^.

To contextualize this to our single nuclei RNA seq analysis, the data segregated into a very small population of myelinating Schwann cells (0.2%, Fig. 2) and a larger population of non-myelinating Schwann cells (5.7%). These clusters shown represent the “typical” Schwann cells based on classic marker expression (*MBP, MPZ, S100B, ERBB3, NCAM1, and SOX10*). We expected the myelinating Schwann cell population to be small as this phenotype is consistently downregulated after transection^13,15,34^. Likely the myelinating and non-myelinating Schwann cells transformed into repair cells after transection^42,44,53^. However, our data suggests that repair Schwann cells may be more heterogenous and widespread in different clusters than the clearly-defined Schwann cell clusters shown.

Many clusters beyond the defined Schwann cell clusters may contain repair Schwann cells including the heterogenous, mesenchymal, and epineurial mesenchymal cell clusters. We observed in these clusters the expression of *NGFR* (a marker for Schwann-like glia cells) that may represent the presence of repair Schwann cells^35^. *NGFR* is highly expressed during development, but expression goes down when the axon is mature^56^. *NGFR* is re-expressed in Schwann cells when there is axon or myelination degeneration^56^. The heterogenous cell cluster in our analysis shows the most NGFR-positivity only second to the non-myelinating Schwann cell cluster, thus we have interpreted that this cluster may contain repair Schwann cells. We have named this as a heterogenous cluster due to the presence of other cell types like perineurial cells and mesenchymal cells (Fig 2C). Further histology would be needed to confirm the phenotypes.

As for other markers that typically characterize repair Schwann cells, they were present in the Schwann and in the heterogenous cell clusters, but do not segregate as their own defined cluster in the UMAP plot. These markers are *NCAM1, NGFR*, and *SOX2* for immature Schwann cells. Mature Schwann cells de-differentiate into a flexible phenotype after injury. Other markers of repair Schwann cells expressed in our reparative peripheral nerve tissue were *EGR2, SHH*, but the presence of these markers were not very high in the tissue. We had expected that Schwann cells would be a bigger proportion of cells in our results^37^. The lower Schwann cell count could be explained by the mature Schwann cells having already de-differentiated into a mesenchymal cell type at 2 weeks.

The connection between mesenchymal cells and Schwann cells should not be overlooked. Clements et al. revealed novel aspects of Schwann cell de-differentiation after nerve transection including the transformation into a mesenchymal phenotype^34^. They found that TGF-beta reprograms the bridge Schwann cells, involved in reconnecting the axon into mesenchymal-like cells and a migratory phenotype to drive cells across the wound^34^. Our data revealed sizable mesenchymal cell populations (mesenchymal cluster, endoneurial mesenchymal cluster) possibly due to the de-differentiation of Schwann cells after transection. Further, a marker for mesenchymal cells, PDGFRA in our data was also found in other clusters like the heterogenous cell cluster suggesting that mesenchymal cells may exist beyond just the clearly defined mesenchymal/epineurial mesenchymal clusters.

#### Macrophages as reparative and anti-inflammatory

Our data show that macrophages were 12.2% of the total cells 2 weeks after transection, and the fourth largest cluster of cells after transection. Macrophages play dichotomous roles in injury in pro-inflammatory (M1) and anti-inflammatory (M2) ways. M1 macrophages are pro-inflammatory and secrete cytokines, and M2 macrophages are anti-inflammatory and contribute to tissue repair^57^. The switch in the polarization of their phenotype is influenced by their environment. The dual roles of macrophages allows them to contribute in tissue homeostasis such as in injury progression, and also tissue repair. Soon after injury, macrophages also play an important role in engulfing myelin and axonal debris^57^. M1 pro-inflammatory macrophages release chemokine ligand 2 (CCL2), inducible nitric oxide synthase, and tumor necrosis factor (TNF)-related apoptosis-inducing ligand. M1 macrophages promote the removal of debris and clearing of apoptotic cells^58^. The many subtypes of M2 macrophages induce anti-inflammatory effects to promote the resolution of inflammation, cell proliferation, growth factor production, tissue repair, angiogenesis and wound healing^58-60^. Our data did not segregate into M1 and M2 phenotypes, however transplanting the macrophages in the M2 phenotype could be beneficial to degenerating neurons contributing to tissue repair^58-60^.

### Data interpretation

Even as a powerful tool, there are limitations to the single nuclei RNA seq approach^61^. The clusters of cells are generated via automatic bioinformatics analysis and may or may not reveal all the actual cell types as evidenced by anatomical location, function or immunochemistry. One of the clusters exhibited the heterogeneity of several cell types including perineurial cells, and markers for Schwann cells, and mesenchymal cells. More specific histology staining for the characteristic markers of these cell is need to confirm cell identities.

One concern from our previous whole tissue RNA sequencing analysis^13^ had been the time between excision and freezing of the tissue; but in later proteomic analyses, differences in sample freezing times did not appear to account for proteomic differences^15^. Meanwhile, in the two participants whose peripheral nerve tissue underwent single nuclei RNA seq analysis, the UMAP display of cell clusters were concordant even though there was a greater than 30 minute difference (67 vs. 32 minutes) in freezing times for peripheral nerve tissue between participants. Our interpretation is that the profile of the regenerating peripheral nerve tissue is stable under our collection conditions. We recognize that ideally, the freezing time should be reduced further to more definitively maintain the stability of the samples, but based on the current design of the trial and surgery logistics, a shorter freezing time is not practicably possible. Furthermore, the timing of sample collection of reparative peripheral nerve tissue is the actual timing for the peripheral nerve tissue that is implanted in clinical trials, and we detected and measured the concentration of key factors in this product (Fig 5).

Previous work had transplanted embryonic stem cells, and embryonic dopaminergic neurons into the SN of participants with PD^62-64^. The number of cells transplanted have varied across studies, some limited by the availability of embryos and stem cells. The range of experiments is great from transplanting into the putamen 20µl of embryonic mesencephalic tissue containing dopamine neurons from fragments of aborted embryos^2^, to 9,861-21,552 dopaminergic neurons per putamen side^65^, to dopaminergic neurons from 1 or 4 donor embryos per side (approximately 30,000 cells per side for 1 embryo and 70,000-120,000 cells per side from 4 embryos)^1^. Even with fewer peripheral nerve cells transplanted in our current trial, we have observed preliminary results of improved UPDRS part III motors scores at 12 months^9^ and longer durations (unpublished).

We recognize the use of tissue from participants with PD introduces a concern of neuropathies as people with PD have a higher incidence^66,67^. For the single nuclei RNA seq, one of the participants had no history of neuropathy, and one did have a history of neuropathy. Even with these limitations, this study provides insight into the composition of the reparative peripheral nerve tissue implanted in our ongoing clinical trials.

## Summary

In this work, we were able to demonstrate the types of cells and the anti-apoptotic and neuroprotective contents that are transplanted into our participants with PD. Our novel reparative nerve transplantation may also have immediate utility in other neurodegenerative diseases as such as stroke^68-70^, traumatic brain injury (TBI), and Alzheimer’s disease. We found that multiple cell types in reparative peripheral nerve tissue contribute to the production of a wide array of factors utilized in our goal to alter the progression of Parkinson’s Disease.

## Data Availability

All data produced in the present study are available upon reasonable request to the authors. All data produced will be available online for open-access after publication acceptance.

https://uknowledge.uky.edu

## Ethics approval and consent to participate

The University of Kentucky’s Institutional Review Board (IRB) approved the study (clinicaltrials.gov: NCT02369003), and the participants provided written informed consent.

## Clinical Trial

clinicaltrials.gov: NCT02369003

## Acknowledgements

We acknowledge the assistance of the Research Flow Cytometry Core in the Division of Rheumatology at Cincinnati Children’s Hospital Medical Center and Alyssa Sproles. We thank Morgan Yazell for trial execution, Tom Dolan for medical illustration, Dr. Jeremiah Smith for assistance on 10x single cell analysis. Thank you to Drs. Joyce Peng and Noel Chen of Singulomics Corporation. Nuclei isolation, single nucleus RNA sequencing, and analysis were conducted by Singulomics Corporation (https://singulomics.com/, Bronx NY).

